# Atrial fibrillation and mortality in critically ill patients undergoing emergent fixed-wing air ambulance transport: a retrospective cohort study

**DOI:** 10.1101/2024.04.13.24305714

**Authors:** Marcus Fredriksson Sundbom, Hannah Langelotz, Helena Nyström, Roman A’roch, Michael Haney

**Affiliations:** Department of Surgical and Perioperative Sciences, Anesthesiology and Intensive Care Medicine, Umeå University, Umeå, Sweden

**Keywords:** Critical care, Atrial fibrillation, Mortality, Air Ambulances, Epidemiology

## Abstract

**Objective:** To examine if atrial fibrillation at the time of emergency transport to tertiary care hospital by air ambulance is associated with increased mortality risk.

**Design:** Retrospective cohort study. Logistic regression and Cox regression were used to assess associations of condition with outcome (mortality). Propensity score matching (1:1) was used to identify the comparison cases.

**Setting:** One tertiary care centre in sparsely populated northern Sweden

**Participants:** Critical care cases undergoing emergent fixed-wing air transport in the northern healthcare region of Sweden was included. 2143 cases were included in the study between 2000-2016.

**Primary and secondary outcomes measures:** The primary outcome was mortality risk at 90-days post transport in cases with atrial fibrillation during the fixed-wing air ambulance transport. The secondary outcome measures were vasopressor and impaired oxygenation was associated with mortality at 90-days post transport.

**Results:** Atrial fibrillation showed a higher 90-day post transport mortality risk estimate in the multivariate analysis of all cases, hazard ratio (HR) 1.70 (95% CI 1.08-2.67, p=0.022). In a propensity matched analysis, no increased mortality risk for atrial fibrillation was observed at the same time interval with hazard ratio 1.22 (0.65-2.27, p=0.536). In the secondary analysis, vasopressor use was associated with atrial fibrillation at the time of transport whereas impaired oxygenation was not associated with an increased risk of atrial fibrillation.

**Conclusion:** No increased mortality risk at 90 days was seen for cases with atrial fibrillation at time of transport in this cohort using a propensity score matched approach.

**Article summary:** *Strengths and limitations:*

- Explores the scarcely studied topic of mortality risk and atrial fibrillation at time of transport with fixed-wing air ambulance
- Quasi-experimental approach with propensity score matching to handle selection bias
- No co-morbidity and disease severity data which risk confounding results
- A single transport system was studied which limits generalizability

## Introduction

Atrial fibrillation (AFib) is the most common cardiac arrythmia among high income countries and is associated with an increased risk of stroke, heart failure and mortality[1]. Advanced age, genetics and an unhealthy lifestyle can predispose an individual to develop AFib, and patients typically present with palpitations, dyspnea, chest pain and/or near syncope [2].

AFib is common in critically ill patients, and has been reported to be present in more than one fourth of patients at some point during intensive care treatment[3]. It has been suggested as a predictor of increased mortality risk independent of disease severity and co-morbidity [4–8], especially if the AFib is new-onset[9–11]. Some studies suggest AFib is associated with severity of illness but not mortality[12]. Risk factors for developing new-onset AFib include for example patient age, fluid overload, hypoxemia, hypotension, admission diagnosis and vasopressor use[13]. Several strategies to manage AFib in the ICU can be deployed, such as rhythm control, rate control or pharmacological/electrical cardioversion[14]. However, a recent study reported that the most common strategy amongst ICU doctors in hemodynamically stable ICU patients was observation[15].

Air ambulance transport is common for transport of emergency intensive care cases if there are long distances to tertiary care hospitals. Little is reported about how common AFib is in this group of patients. It is not known if AFib at the time of transport is associated with increased later mortality. We hypothesized that AFib would be associated with increased mortality at 90 days in a mixed cohort of critically ill patients undergoing emergency fixed-wing inter-hospital air ambulance transfer over long distance to tertiary care. A secondary hypothesis was that vasopressor use and impaired oxygenation would be associated with increased risk of AFib.

## Material and methods

Intensive care cases that were transported in the northern healthcare region of Sweden by air ambulance between year 2000-2016 were included in the analysis, from a total of 3917 cases, with 2143 meeting the inclusion criteria of being an emergent transport, >16 years of age, not transported for organ transplantation, and not missing mortality data. Patient charts for the air ambulance cohort was abstracted into a database, with patient data pseudonymized. Mortality data was obtained from the Swedish National Board of Health and Welfare and linked to the cases in the database. SpO2/FiO2 (S/F) (blood oxygen measured by pulse oximetry/fraction of inspired oxygen) ratio was used as a surrogate for PaO2 (partial pressure of oxygen in blood)/FiO2[16]. Impaired oxygenation was defined according to ARDS network guidelines[17], with cases with S/F ratio <300 regarded as having impaired oxygenation. Cases were divided into diagnostic groups according to the reason for initiating the transport.

Concerning analysis, summary statistics are presented with mean (range) for continuous variables and counts (percentages) for categorical variables. Logistic regression and Cox regression was used to assess association on mortality of various patient level factors and significance level set to alpha 0.05. Complete case analysis was used. All statistical analysis were made using SPSS 28 (IBM Corporation, Armonk, NY, USA) and forest plots were made using GraphPad Prism 9 (GraphPad Software, San Diego, CA, USA). Propensity score matching 1:1 was done in SPSS based on age, sex, mechanical ventilation, vasopressor, vasodilator, inotrope, oxygenation status and diagnostic group. These factors were either selected based on univariate and multivariate mortality risk assessment or by being clinically relevant factors for increased mortality which been previously reported [18].

## Results

### Study population

2143 cases met the inclusion criteria and were included in the initial analysis (Table 1). Of these, 79 cases presented with AFib. Propensity matching was undertaken, and characteristics of this group is presented in Table 1. Propensity score distribution is presented in Fig 2.

**Table 1.**
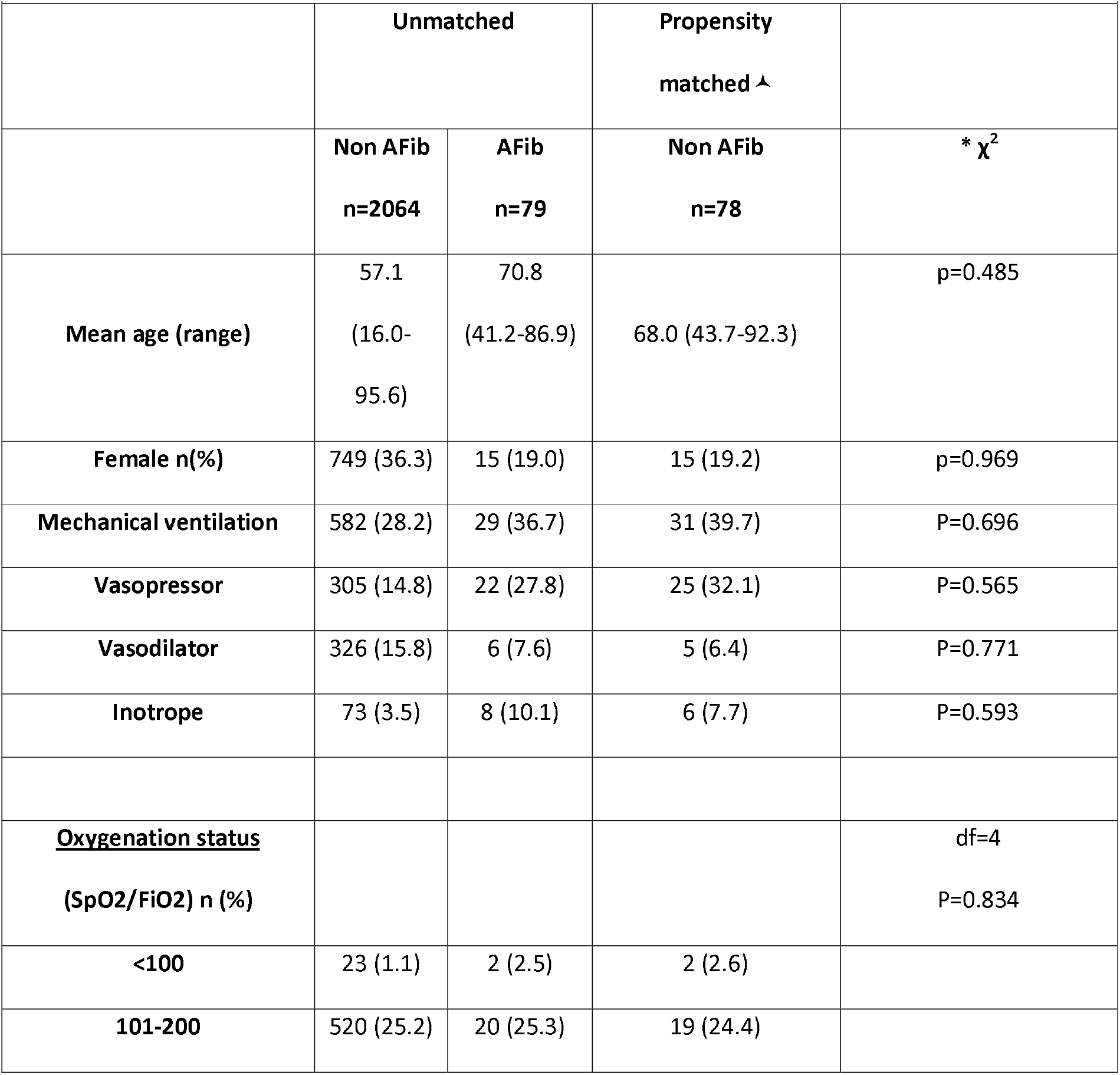
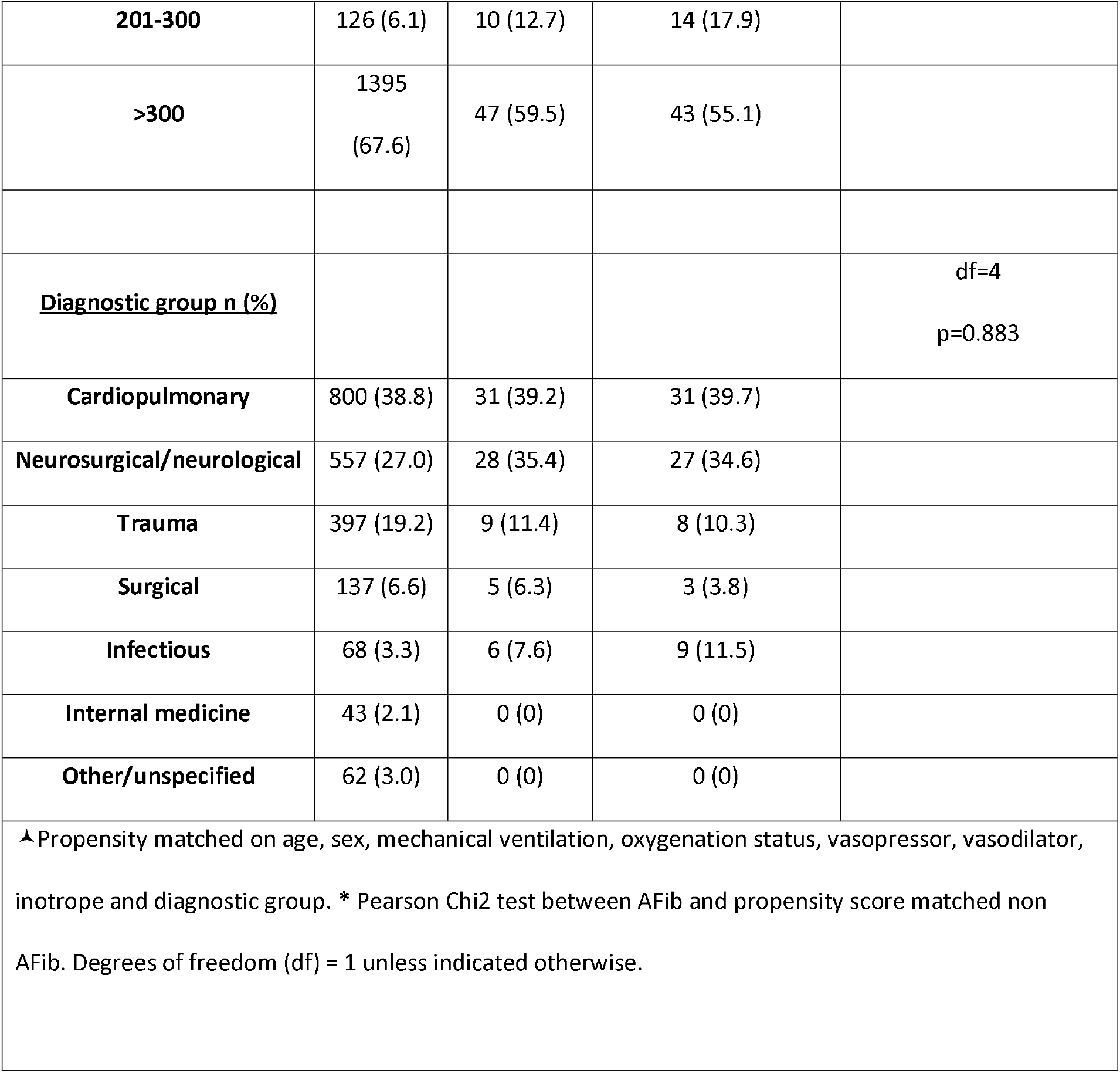
Characteristics of the cohort.

**Fig 1.**
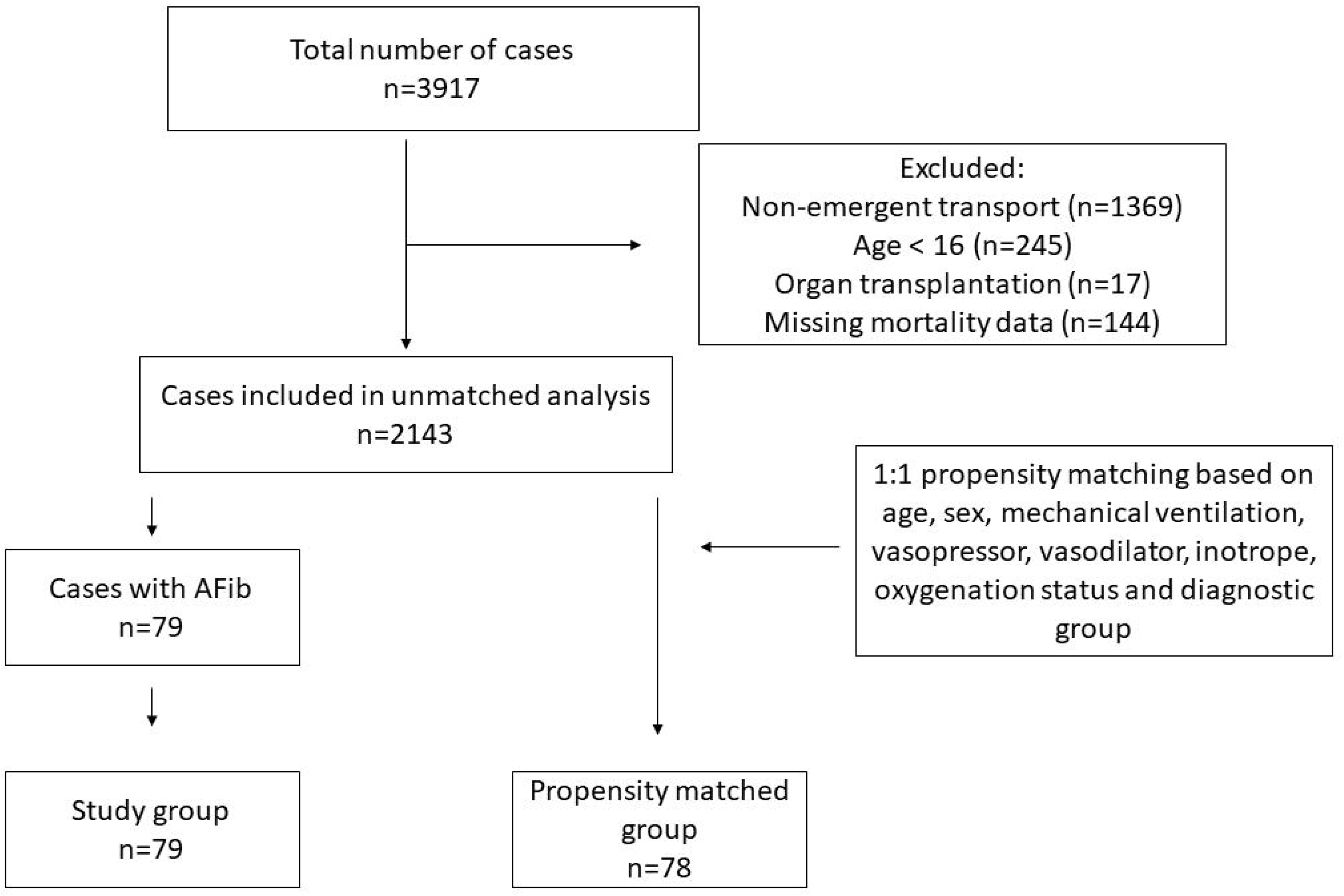
Flowchart for patients included the analysis.

**Fig 2.**
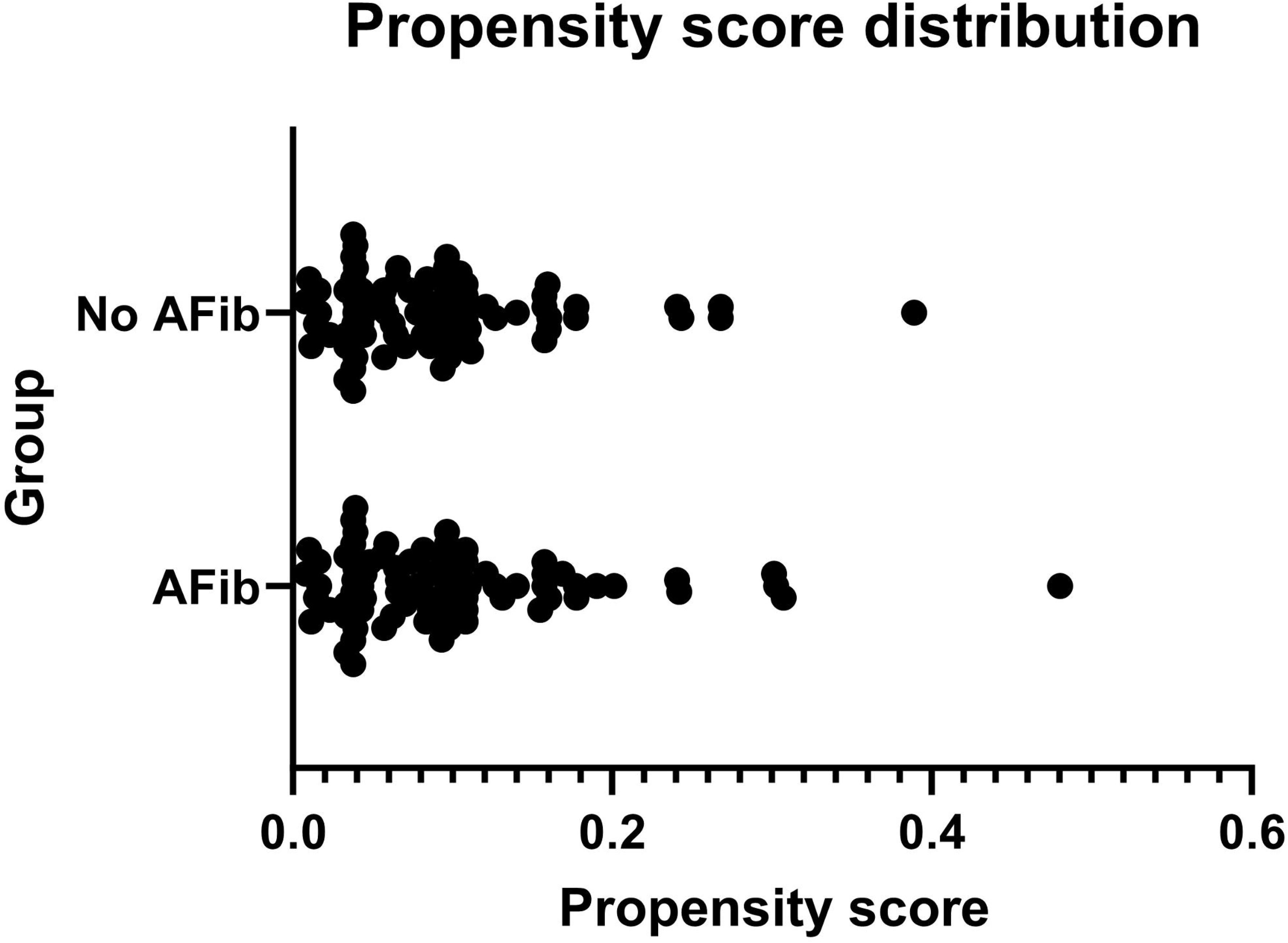
Propensity score distribution. Distribution of propensity score across the AFib and non AFib group

### Mortality risk

An increased mortality risk for the AFib group at 90 days post transport was observed compared to non-AFib cases in the unmatched cohort (Fig S1), but could not be observed in the propensity-matched analysis using Cox regression, hazard ratio 1.22 (95% confidence interval (CI) 0.65-2.27, p=0.536).

### Risk of atrial fibrillation

When other conditions were analyzed in the unmatched cohort, the use of a vasopressor or inotrope was associated with an increased risk of AFib with odds ratio 2.09 (95% CI 1.23-3.53, p=0.006) and 2.82 (95% CI 1.26-6.31, p=0.012) respectively (Fig 3.). Age and sex were also associated with an increased risk of AFib. Impaired oxygenation was not associated to an increased risk of AFib (Fig 3).

**Fig 3.**
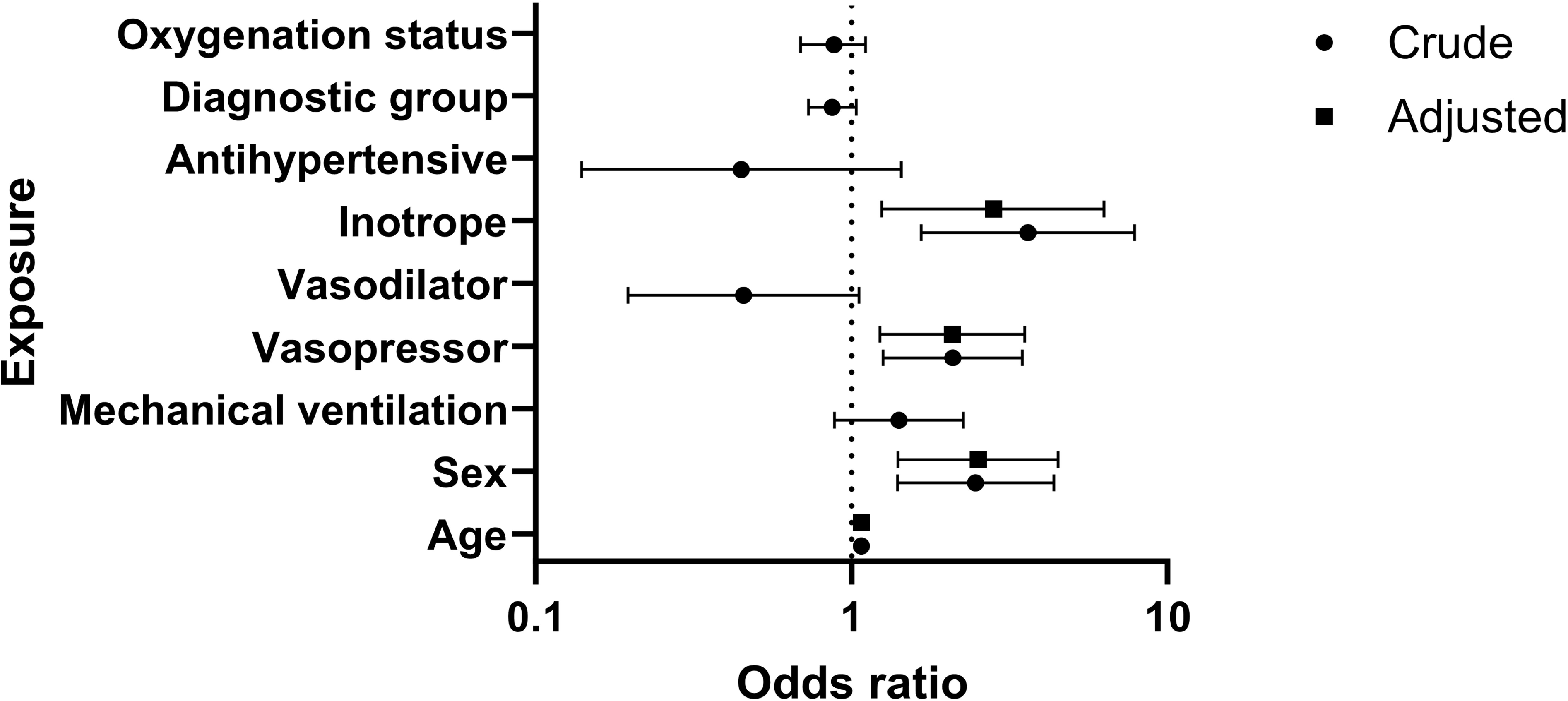
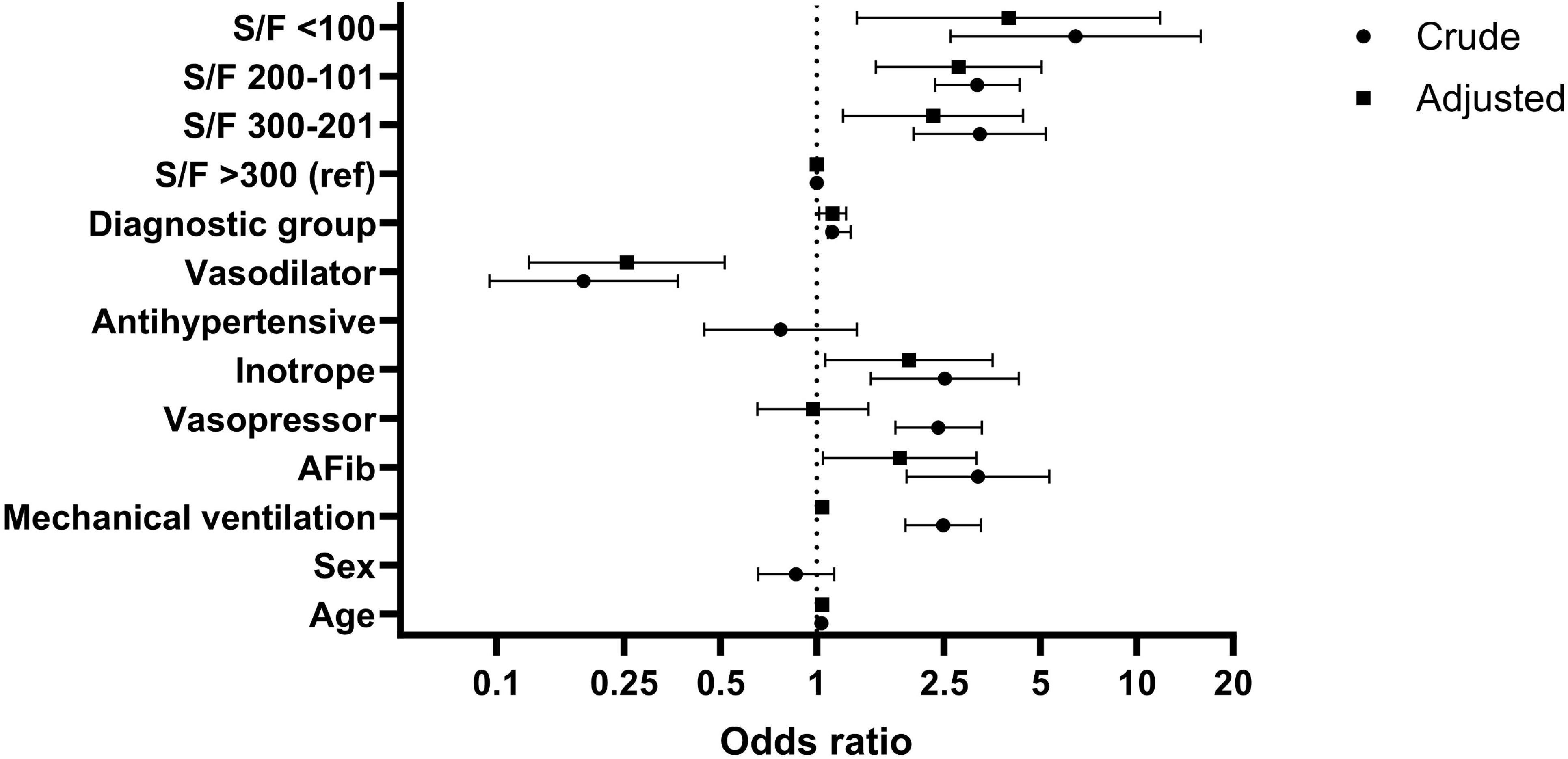
Logistic regression of exposures and AFib risk. Crude represents univariate analysis whereas adjusted represents multivariate analysis with all included exposures

## Discussion

In this study we found that AFib was not associated with increased mortality risk at 90 days using a propensity score matched approach. Several other studies of mixed cohorts of critically ill patients who report no increased mortality risk are in line with our findings [19–22]. A recent study focused on critically ill patients suffering from acute ischemic stroke found an increased crude ICU and in-hospital mortality risk which was not observed after propensity score matching[23]. Increased mortality risk for AFib is however frequently reported in critically ill patients with sepsis or post cardiothoracic surgery[4,10,24,25]. The prevalence of AFib was similar to what has been previously reported in mixed patient ICU cohorts[26]. Increased use of vasopressor and use of inotropic drugs has previously been associated with increased AFib risk which we also observed [27–29]. There was no association between impaired oxygenation and AFib risk. Although an increased risk of AFib cases with impaired oxygenation has been previously reported, most recently in cases with severe COVID-19 [30–32], many patients (37%) in our cohort (pre-pandemic) had impaired oxygenation which may in part explain this disparity. Furthermore, the duration of impaired oxygenation may be different compared to other studies, but this cannot be assessed.

### Limitations

This study had several limitations. First, there was no access to pre-ICU transport co-morbidity data. As a result, there could be adjustment only for the diagnosis which was the reason for initiating the transport. Second, conclusions about the nature of the AFib could not be drawn since information about pre-existing conditions was unavailable to us. Third, only one large regional transport system cases were studied, which limits generalizability of the findings. Another factor is that the cases in this study had all been pre-stabilized at sending hospital, and presumably resuscitation with some fluid therapy was initiated before the transport to treat hypovolemia. Fluid overload is associated with increased risk of AFib[33,34], but to assess to which extent the cases had received appropriate fluid therapy was beyond the scope of this study.

## Conclusion

This study was able to analyze AFib-associated factors in the context of critical care transport by air ambulance in a regional cohort. AFib at the time of transport was not associated with increased mortality risk in this cohort. Risk of AFib was associated with use of vasopressor but not associated to impaired oxygenation, but no causal inference can be drawn from this analysis.

## Author contributions

MFS primary responsible as guarantor. RA and MH was responsible for data collection and curation. MFS, HL, MH, HN, RA conceptualization, analysis, interpretation and original draft.

## Acknowledgements

We acknowledge the contribution of Göran Johansson for his role in data curation.

## Data availability

Data from which the study is derived are available upon reasonable request from the corresponding author. Data is not made immediately accessible because of Swedish privacy laws.

## Ethics statements

### Patient consent for publication

Not applicable

### Ethical approval

Ethical permission was granted by the Swedish Ethical Review Authority (Dnr 2019-06525).

